# Robust ACE2 protein expression localizes to the motile cilia of the respiratory tract epithelia and is not increased by ACE inhibitors or angiotensin receptor blockers

**DOI:** 10.1101/2020.05.08.20092866

**Authors:** Ivan T. Lee, Tsuguhisa Nakayama, Chien-Ting Wu, Yury Goltsev, Sizun Jiang, Phillip A. Gall, Chun-Kang Liao, Liang-Chun Shih, Christian M. Schürch, David R. McIlwain, Pauline Chu, Nicole A. Borchard, David Zarabanda, Sachi S. Dholakia, Angela Yang, Dayoung Kim, Tomoharu Kanie, Chia-Der Lin, Ming-Hsui Tsai, Katie M. Phillips, Raymond Kim, Jonathan B. Overdevest, Matthew A. Tyler, Carol H. Yan, Chih-Feng Lin, Yi-Tsen Lin, Da-Tian Bau, Gregory J. Tsay, Zara M. Patel, Yung-An Tsou, Chih-Jaan Tai, Te-Huei Yeh, Peter H. Hwang, Garry P. Nolan, Jayakar V. Nayak, Peter K. Jackson

**Affiliations:** Department of Pathology, Stanford University School of Medicine, Stanford, California 94305, USA; Department of Otolaryngology–Head and Neck Surgery, Stanford University School of Medicine, 801 Welch Road, Stanford, CA, USA; Division of Allergy, Immunology, and Rheumatology, Department of Pediatrics, Stanford University School of Medicine, Stanford, CA, USA; Baxter Laboratory, Department of Microbiology & Immunology, Stanford University School of Medicine, Stanford, California, USA; Department of Otolaryngology, National Taiwan University Hospital, Taipei, Taiwan; Department of Otorhinolaryngology, China Medical University Hospital, Taichung, Taiwan; Graduate Institute of Biomedical Sciences, China Medical University, Taichung, Taiwan; Terry Fox Cancer Research Laboratory, Translational Medicine Center, China Medical University Hospital, Taichung, Taiwan; School of Medicine, China Medical University, Taichung, Taiwan; Department of Otolaryngology–Head and Neck Surgery, Columbia University School of Medicine, New York City, NY; Department of Otolaryngology–Head and Neck Surgery, University of Minnesota School of Medicine, Minneapolis, MN; Department of Otolaryngology–Head and Neck Surgery, University of California San Diego School of Medicine, San Diego, CA; Division of Immunology and Rheumatology, Department of Internal Medicine, China Medical University Hospital, Taichung, Taiwan

## Abstract

We investigated the expression and subcellular localization of the SARS-CoV-2 receptor, angiotensin-converting enzyme 2 (ACE2), within the upper (nasal) and lower (pulmonary) respiratory tracts of healthy human donors. We detected ACE2 protein expression within the cilia organelle of ciliated airway epithelial cells, which likely represents the initial or early subcellular site of SARS-CoV-2 viral entry during respiratory transmission. We further determined whether ACE2 expression in the cilia of upper respiratory cells was influenced by patient demographics, clinical characteristics, co-morbidities, or medication use, and found no evidence that the use of angiotensin-converting enzyme inhibitors (ACEI) or angiotensin II receptor blockers (ARBs) increases ACE2 protein expression.

---

Coronavirus disease 2019 (COVID-19) is an ongoing pandemic infection caused by the positive-sense RNA virus, severe acute respiratory syndrome coronavirus 2 (SARS-CoV-2)^1^. The high transmissibility of the virus, along with case fatality estimates ranging from 1% to above 5%, has raised concerns worldwide. Patients with comorbid conditions including hypertension, diabetes, and pulmonary disease are highly represented among hospitalized patients with COVID-19 disease, suggesting the presence of risk factors that may determine susceptibility to SARS-CoV-2 infection ^2–5^

A molecular connection between SARS-CoV-2 and hypertension, in particular, is suggested by the discovery that ACE2 is the major essential receptor for SARS-CoV-2 ^6,7^. ACE2 plays an important role in the renin-angiotensin-aldosterone system (RAAS), which consists of a cascade of vasoactive peptides that maintain blood pressure and electrolyte homeostasis. ACE2 converts vasoconstrictor peptides, angiotensin (Ang) II and Ang I, into the vasodilator peptides, Ang (1-7) and Ang (1-9), respectively ^8^. These actions counterbalance the enzymatic effect of the related ACE, which generates angiotensin II from angiotensin I. ACEI and ARBs are commonly used antihypertensive drugs that target components of the RAAS. Several recent correspondences have raised concerns that ACEI and ARBs may increase expression of ACE2 and thereby elevate the risk of infection by SARS-CoV-2, thus potentially explaining why hypertension is a common comorbidity in patients with COVID-19 ^9–12^. This hypothesis is also rooted in human and rodent studies showing upregulation of *ACE2* mRNA in the heart, kidney, and urine after ACEI/ARB administration^13–15^. Notably, however, the effects of ACEI and ARBs on the expression of ACE2 in the respiratory tract are currently unknown. Given SARS-CoV-2 causes respiratory infections, whether ACE2 expression is altered in the airway of patients taking ACEI or ARBs is a critical question that needs to be addressed to support continued clinical use of these antihypertensive drugs.

We first determined the expression patterns of the ACE2 protein in the upper and lower respiratory tract. Gene expression analyses have identified *ACE2* expression in the nasopharynx, oral mucosa, lungs, intestines, kidney, and testis^16^, and protein expression studies have largely been concordant with these tissue-specific findings^17,18^. However, a recent preprint suggests the absence of the ACE2 protein in the lung, bronchus, and nasopharynx^19^. In order to understand the precise nature of ACE2 protein expression in tissues relevant for COVID-19, we performed immunohistochemistry using a panel of ACE2 antibodies on human tissues. Consistent with prior studies, we found that several ACE2 antibodies appropriately stain ACE2 in the kidney, testis, seminal vesicles, and intestinal villi (Extended Data Fig. 1). However, only two antibodies that we tested (Abcam ab15348 and Sigma HPA000288), stained ACE2 in the CD31+ vascular endothelium (Extended Data Fig. 2a), where ACE2 expression has been reported^17^. In the lungs, anti-ACE2 clone (Abcam ab15348) yielded robust staining of pneumocytes, while the other clones showed only weaker or negligible signal (Extended Data Fig. 1 and 2b). After careful antibody titration, clone selection, and validation across multiple tissue types, we find that the overall expression intensity of ACE2 in the lung is low when compared to the kidney, testis, and intestinal villi (Extended Data Fig. 1).

**Extended Data Fig. 1.**
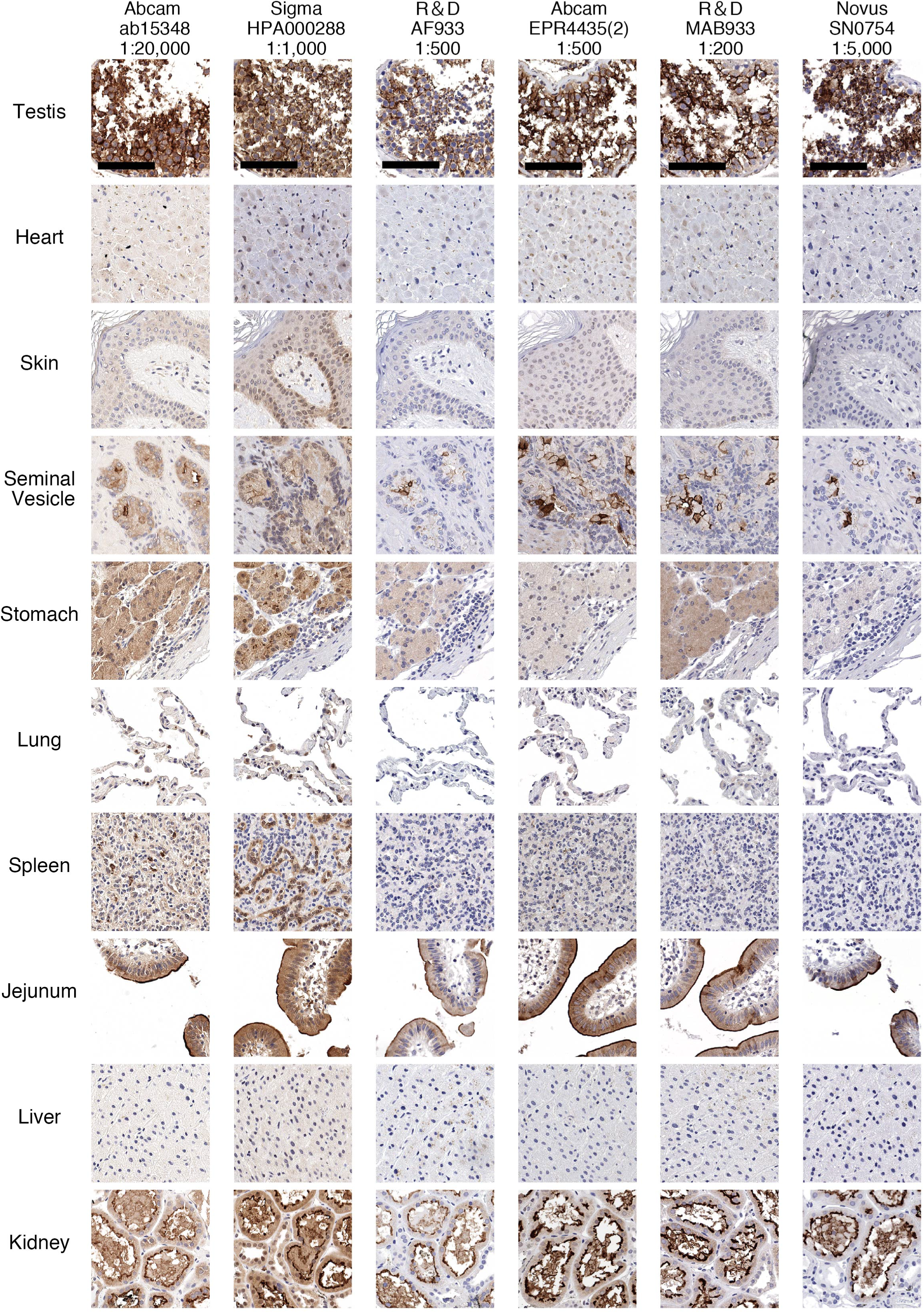
Immunohistochemical analysis of ACE2 protein localization across human tissues using multiple anti-ACE2 antibodies. Representative images of human tissues stained by chromogenic immunohistochemistry using antibodies targeting the ACE2 protein (brown) and counterstained with hematoxylin (blue). Highest ACE2 expression was observed in the villi of the intestinal tract (jejunum), renal tubules, testis, and glandular cells in the seminal vesicle. Minimal to no/non-specific staining can be seen in the heart, stomach, spleen, skin, and liver. Staining of lung pneumocytes was observed using Abcam ab15348 and Sigma HPA000288 (also see Extended Data Fig. 2b). Scale bars: 100 μm.

**Extended Data Fig. 2.**
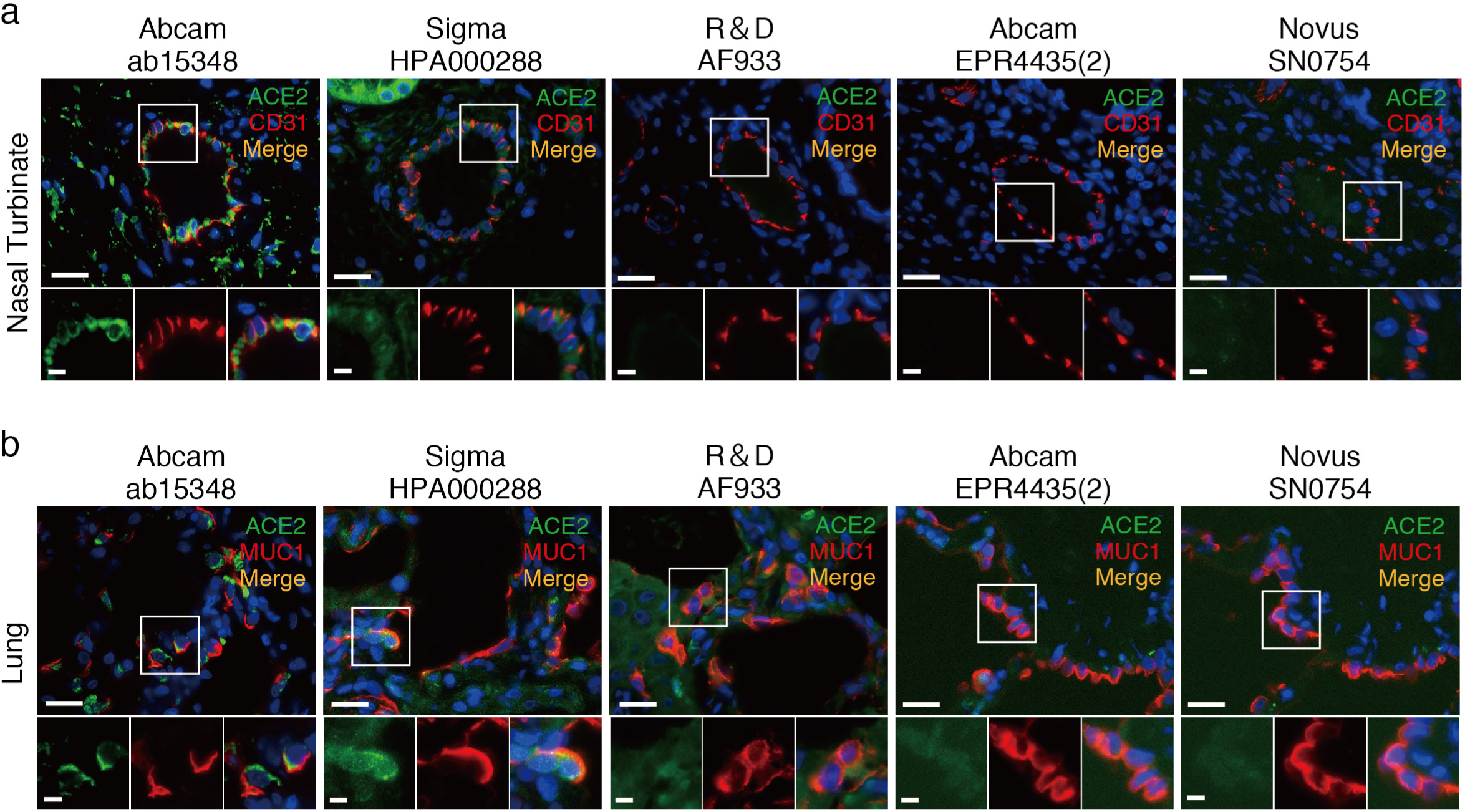
ACE2 protein expression within human vascular endothelial cells and the lung. a, Representative double immunofluorescence staining of ACE2 and endothelial cell marker, CD31 in the blood vessel of human nasal turbinate using 5 different anti-ACE2 antibodies and anti-CD31. b, Double immunofluorescence staining of ACE2 and type II pneumocyte marker, mucin 1 (MUC1) in the human lung using 5 different anti-ACE2 antibodies and an anti-MUC1 antibody. The scale bars are 20 μm (top) and 5 μm (bottom).

We performed double immunofluorescent staining of ACE2 with mucin 1 (MUC1), an established type II pneumocyte marker, and confirmed that of tested antibodies, Abcam ab15348 had the most specific staining patterns and showed that ACE2 is expressed within type II pneumocytes of human lung (Extended Data Fig. 2b). These findings support recent single-cell RNA-sequencing (scRNA-seq) data showing *ACE2* enrichment within type II pneumocytes^20,21^. Our results overall support the specificity of some commercially available antibodies by orthogonal validation and validate the presence of ACE2 protein within the human airway. These antibody testing results may also serve as a useful resource to help guide future protein-based studies.

We next investigated ACE2 protein expression within the epithelium of the human respiratory tract using anti-ACE2 (ab15348) given its robust and specific staining patterns. Recent studies using scRNA-seq have identified *ACE2 mRNA* expression within ciliated epithelial cells in the nasal cavity^20–22^. Sungnak et al. demonstrated that ciliated nasal epithelial cells have one of the highest *ACE2* mRNA expression levels among investigated cell types in the respiratory tract^21^.

We therefore performed double immunofluorescence staining using anti-ACE2 and anti-acetylated α-tubulin (ACTUB), a marker of the cilia organelle, and discovered that not only is ACE2 expressed in ciliated epithelial cells, but within these cell types, ACE2 is abundantly present in the cilia organelle of human nasal turbinate, ethmoid sinus, uncinate process (sinus), trachea, and bronchial epithelial cells (Fig. 1a) compared to appropriate isotype controls (Extended Data Fig. 3). We also examined the mouse respiratory tract and found that ACE2 is similarly expressed within the cilia of the mouse trachea and nasal turbinate (Fig. 1b). Staining of ACE2 within IMCD3 cells, a ciliated kidney epithelial cell line, further confirms ACE2 localization within renal primary cilia (Fig. 1c). Overexpression of human *ACE2* in IMCD3 cells showed a predicted increase in the percentage of ACE2 staining in the primary cilia (Fig. 1d, e), providing additional evidence of ACE2 localization to the primary cilia and further validation of the antibody’s specificity. These findings are consistent with a prior study showing ACE2 protein co-localization with β-tubulin IV, another cilia marker, from human airway epithelial cells cultured at the air-liquid interface^23^. Importantly, both the transcript and protein for transmembrane serine protease 2 (TMPRSS2), a serine protease required for the crucial spike protein activation step of SARS-CoV-2 entry, is also expressed in the cilia organelle of the human respiratory tract,^18,20,24^.

**Fig. 1.**
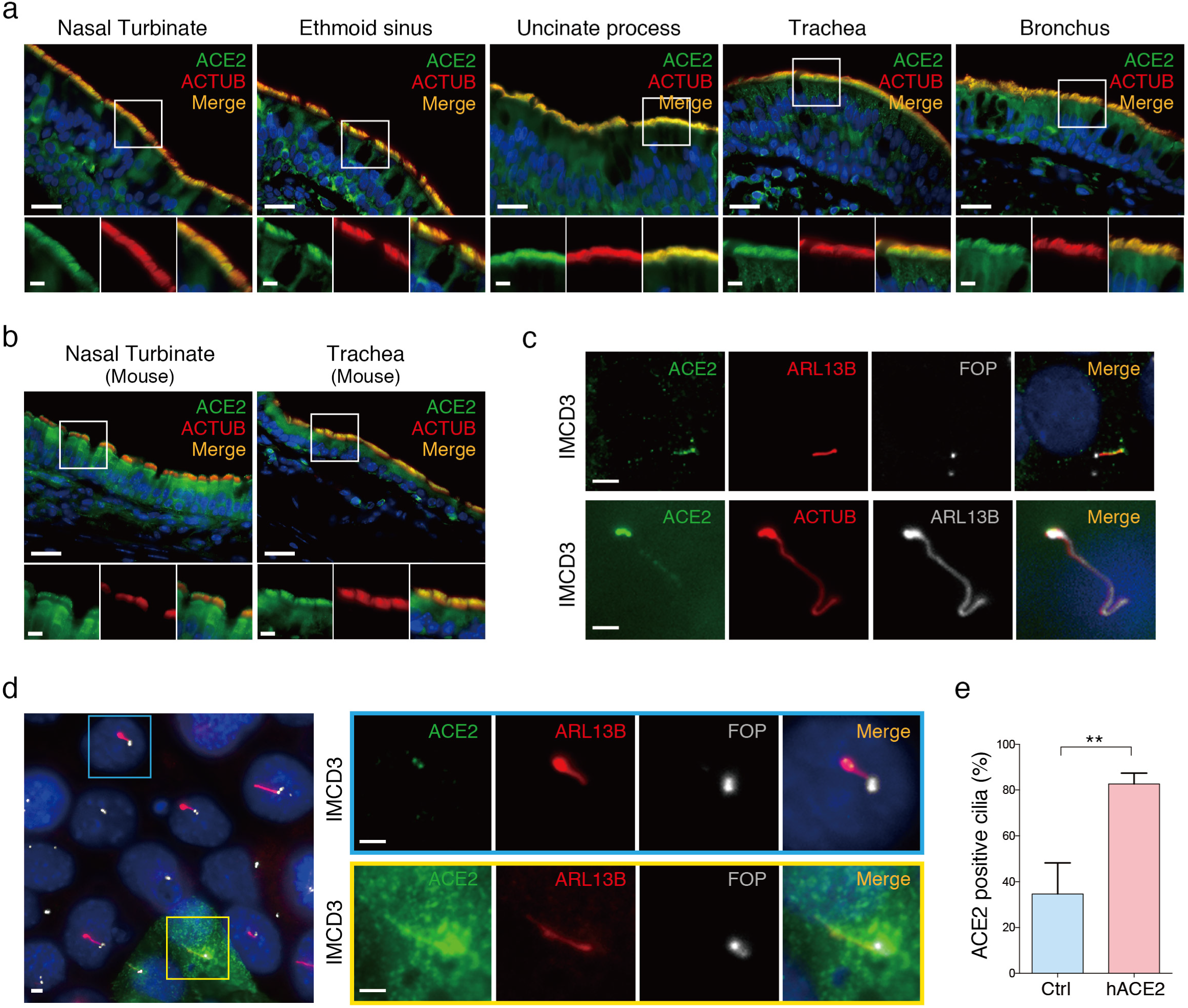
ACE2 protein expression in the cilia organelle of ciliated epithelial cells in the upper and lower respiratory tract and in a ciliated kidney epithelial cell line. **a**, Representative double immunofluorescence staining of ACE2 and acetylated α-tubulin (ACTUB) on normal human nasal turbinate, ethmoid sinus, uncinate process (sinus), trachea, and bronchus, using anti-ACE2 and anti-ACTUB antibodies, respectively. **b**, Representative double immunofluorescence staining of ACE2 and ACTUB on normal C57BL/6J mouse nasal turbinate and trachea. **c**, Immunofluorescent staining of (top panel) ACE2, cilia marker ADP-ribosylation factor-like protein 13B (ARL13B), and cilia centrosome marker FGFR1 Oncogene Partner (FOP); (bottom panel) ACE2, and cilia markers ACTUB and ARL13B in a ciliated mouse cell line, IMCD3. **d**, Immunofluorescent staining of ACE2 in the primary cilia of IMCD3 cells transiently transfected with human *ACE2* (yellow outline) compared to endogenous mouse ACE2 (blue outline). **e**, Quantified percentages of endogenous ACE2-positive cilia (34.67 ± 13.58%; control (Ctrl)) versus cilia with overexpressed human *ACE2* (82.67 ± 4.73%). Ciliated cells were identified by staining of ARL13B. Error bars represent mean ± SD. (n = 3 independent experiments with 100 cells scored per experiment). (Student’s t test, **p<0.01). The nuclei were stained using DAPI (blue) as a counterstain. Scale bars: 20 μm (**a,b**, top); 5 μm (**a,b**, bottom); 2 μm (**c,d**).

**Extended Data Fig. 3.**
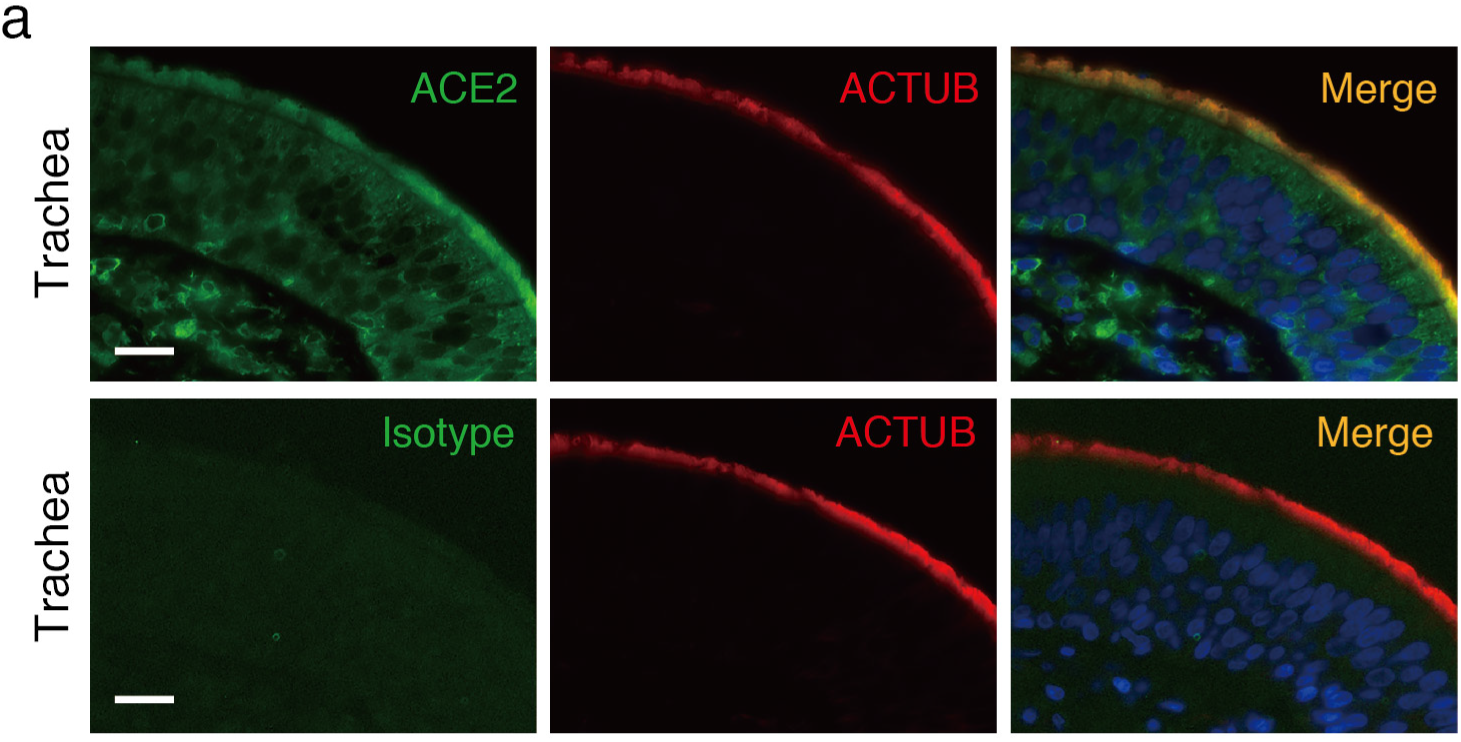
ACE2 expression compared to isotype control in human tracheal tissue. Representative immunofluorescence double staining of ACE2 and ACTUB using anti-ACE2 and anti-ACTUB, respectively, in the top panels compared to double staining of isotype control and ACTUB using rabbit IgG isotype and anti-ACTUB, respectively, in the bottom panels. The nuclei were stained using DAPI. The scale bars are 20 μm.

Taken together, our data suggest that the respiratory tract cilia organelle contains the necessary molecular components for SARS-CoV-2 entry. Indeed, ciliated nasal epithelial cells have been known to be early sites of viral contact for many respiratory viruses including SARS-CoV^25^, MERS-CoV^26^, rhinovirus^27^, parainfluenza virus^28^, respiratory syncytial virus (RSV)^29^, and influenza^30^, which each exploit respective host proteins harbored within the ciliated nasal epithelial cells. CX3CR1, the receptor for RSV, in particular, is expressed in the respiratory cilia organelle within epithelial cells^31^. The related SARS-CoV has also been localized to the cilia body of the airway^25^. Although future imaging is needed to localize the SARS-CoV-2 virus in the cilia of infected human respiratory tissues, our data strongly suggests that ACE2 protein in the cilia body of the respiratory tract, represents the probable initial or early subcellular site of SARS-CoV-2 viral entry.

Curiously, using double immunofluorescent staining of ACE2 with MUC5AC, a goblet cell marker, we find no evidence of ACE2 protein expression within the non-ciliated goblet cells of the respiratory tract (Extended Data Fig. 4a). Since this result differs from scRNA-seq findings of *ACE2* mRNA expression within goblet cells of nasal turbinates and ethmoid tissue from healthy donors and patients with chronic rhinosinusitis (CRS)^20,21^, we then performed *in situ* hybridization using an *ACE2* probe in combination with an anti-MUC5AC antibody, and found no evidence of *ACE2* mRNA expression within goblet cells of the respiratory tract (Extended Data Fig. 4b). These results may reflect limitations in functional interpretation of current single-cell transcriptomic studies, as well as the importance of targeted transcript and protein validation methods to complement shotgun analytic approaches. In summary, we find no evidence of ACE2 protein nor mRNA expression in goblet cells of the respiratory airway, suggesting that unlike ciliated epithelial cells, goblet cells are unlikely to be directly infected by SARS-CoV-2.

**Extended Data Fig. 4.**
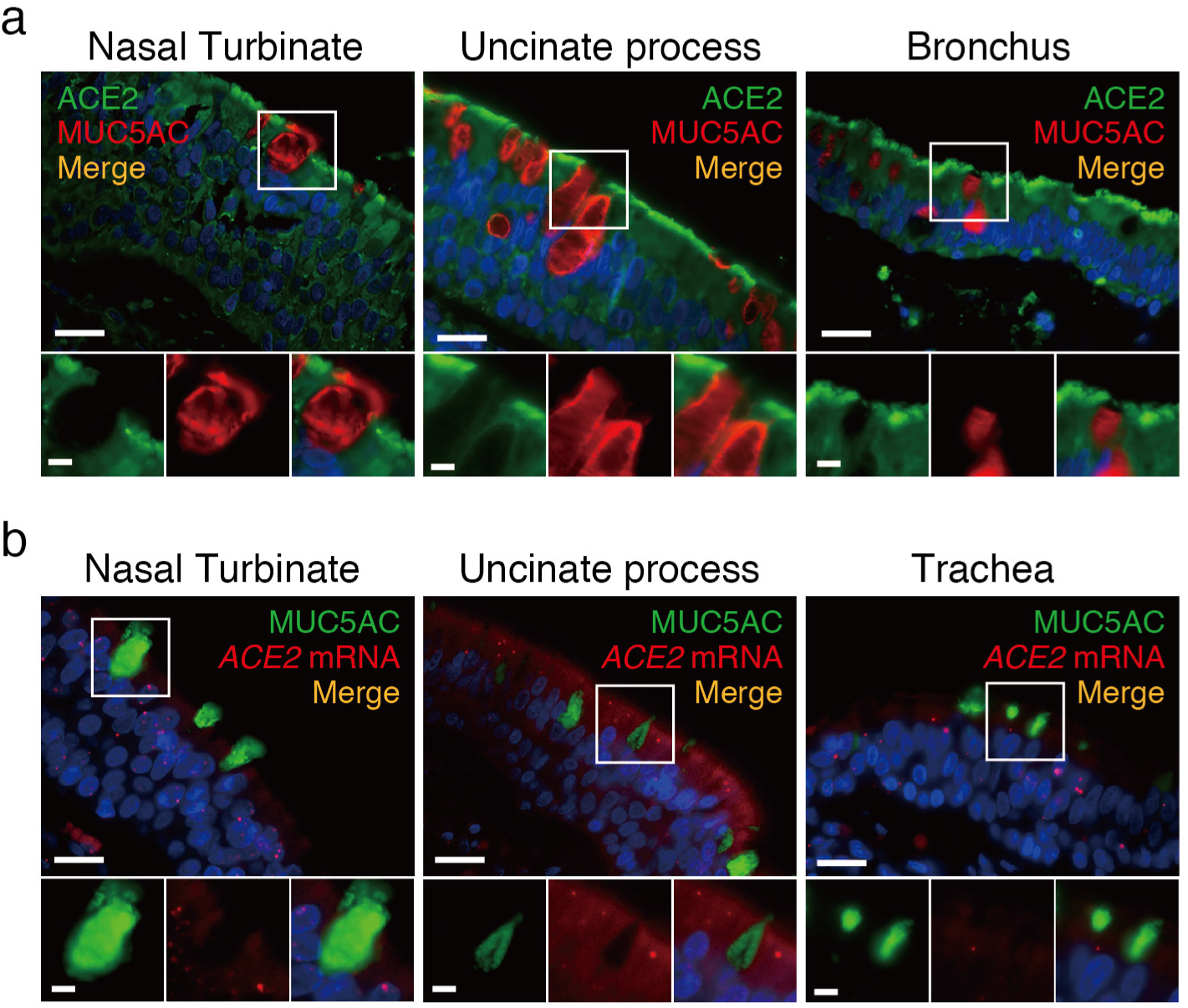
ACE2 mRNA and protein expression are not found in goblet cells of the respiratory tract. **a**, Representative immunofluorescence double staining of ACE2 and mucin 5AC (MUC5AC) reveals the absence of co-localization with goblet cells in the human nasal turbinate, uncinate process, and bronchus. **b**, Representative in situ hybridization using an *ACE2* probe in combination with an anti-MUC5AC antibody. *ACE2* mRNA expression (red dots) was not noted within goblet cells marked by MUC5AC in the nasal turbinate, uncinate process, and trachea. The nuclei were stained using DAPI. The scale bars are 20 μm (top) and 5 μm (bottom).

We next identified patient factors that may contribute to changes in the expression of ACE2 in the nasal epithelial cilia, as this may have important clinical implications for susceptibility to SARS-CoV-2 transmission. Higher ACE2 expression is correlated with higher pseudotype SARS-CoV-2 and SARS-CoV viral infectivity, suggesting that increased ACE2 levels may predispose individuals to SARS-CoV-2 transmission^32–34^. We leveraged our existing human nasal tissue bank, which contains detailed demographics, medical, social, and medication history from patients who have donated their upper airway tissues from 3 academic medical centers (Stanford University, National Taiwan University (NTU), and China Medical University (CMU)) from 2018-2020, to characterize whether ACE2 expression in the upper respiratory cilia is affected by specific patient characteristics.

We determined the extent to which ACE2 expression may differ by age, sex, and smoking status -- three covariates that have been associated with COVID-19 disease severity. Across all three sample cohorts, we found no significant differences in ACE2 expression based on age (≥65 years), sex, or smoking status (Extended Data Fig. 5a). These results differ from some, but not all, recent gene expression studies comparing *ACE2* expression in patients with varying demographics and smoking status^35–37^. Our results suggest that host factors outside of ACE2 expression may determine why males, patients of older age, and smokers are epidemiologically linked to COVID-19 susceptibility.

**Extended Data Fig. 5.**
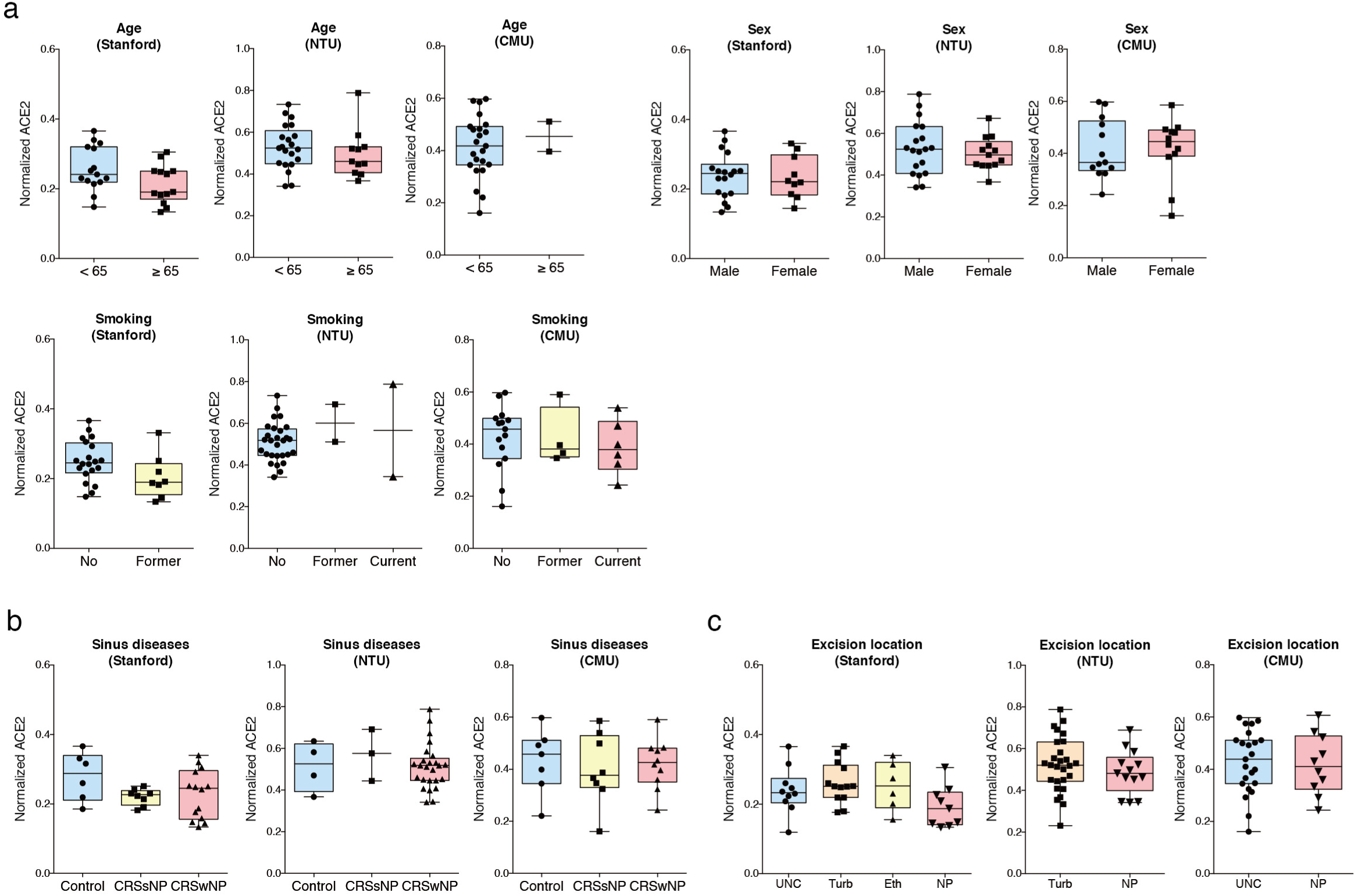
Comparison of ciliary ACE2 protein expression by age, sex, smoking status, sinus disease, and anatomical region. **a**, No statistical significance for ACE2 expression was seen among patients less than or greater than 65 years of age, males versus females, and patients with varying smoking history. (Student’s t-test or Kruskal-Wallis test, p> 0.05). **b**, No statistical significance of ACE2 expression was seen between healthy controls and patients with chronic rhinosinusitis with polyps (CRSwNP) or without polyps (CRSsNP). (Kruskal-Wallis test, p> 0.05). **c**, No statistical significance of ACE2 expression was seen between distinct human nasal tissue sites/regions. (Kruskal-Wallis test, p> 0.05). UNC, uncinate process; Turb, nasal turbinates; Eth, ethmoid sinus; NP, benign nasal polyps.

We next examined whether ACE2 expression in the upper respiratory cilia differs between healthy donors versus patients with chronic rhinosinusitis (CRS), a non-malignant chronic inflammatory disease of the paranasal sinuses that presents either with benign nasal polyps (CRSwNP) or without nasal polyps (CRSsNP). Across all three patient cohorts, no significant differences were noted between healthy donors and patients with chronic rhinosinusitis with or without nasal polyps (Extended Data Fig. 5b). There were also no observed differences in ACE2 expression between anatomical regions within the nasal cavity (Extended Data Fig. 5c). These results suggest that patients with CRS may not be at a higher risk of SARS-CoV-2 infection. There are currently no epidemiological studies that have assessed the prevalence of COVID-19 in patients with CRS to our knowledge.

Finally, we identified patients within our nasal tissue bank who have been taking either ACEI or ARBs for at least six continuous months prior to nasal surgery and compared their ACE2 expression to controls matched for age, sex, and smoking status who have never taken ACEI/ARBs. ACE2 expression was slightly but statistically significantly decreased in patients taking ACEI compared to matched controls in the Stanford cohort, whereas ACE2 expression was not significantly different in patients taking ARBs within all three patient cohorts (Figure 2). We were unable to identify patients taking ACEI in the two Taiwanese cohorts, likely because ARBs are strongly preferred over ACEI for management of hypertension in Taiwan^38^. Subgroup analysis comparing ACEI and ARBs treatment groups to controls of similar age, sex, hypertension, or smoking status revealed a similar trend of lower ACE2 expression in the ACEI and ARB group among most cohort groups, although statistical significance was not attained likely due to the low sample size (Extended Data Fig. 6a,c,d,e). When patients from all three cohorts were combined as a normalized Z-score, not surprisingly, ACE2 expression in the patients taking ACEI was significantly lower compared to controls (Extended Data Fig.6b). Above all, these results suggest that the use of ACEI or ARBs does <u>not</u> increase ACE2 expression in the upper respiratory cilia, and therefore patients on ACEI or ARBs are likely at no greater risk of SARS-CoV-2 transmission than other individuals. Indeed, a recent retrospective cohort study found that the use of ACEI/ARBs was associated with lower risk of all-cause mortality compared with ACEI/ARBs non-users^39^. Although we saw a trend towards decreased ACE2 expression in patients taking ACEI, we strongly caution against interpretation of this result in isolation as suggesting that ACEI is protective against SARS-CoV-2 transmission given our limited sample size, and the likelihood of other host factors that determine viral susceptibility.

**Fig. 2.**
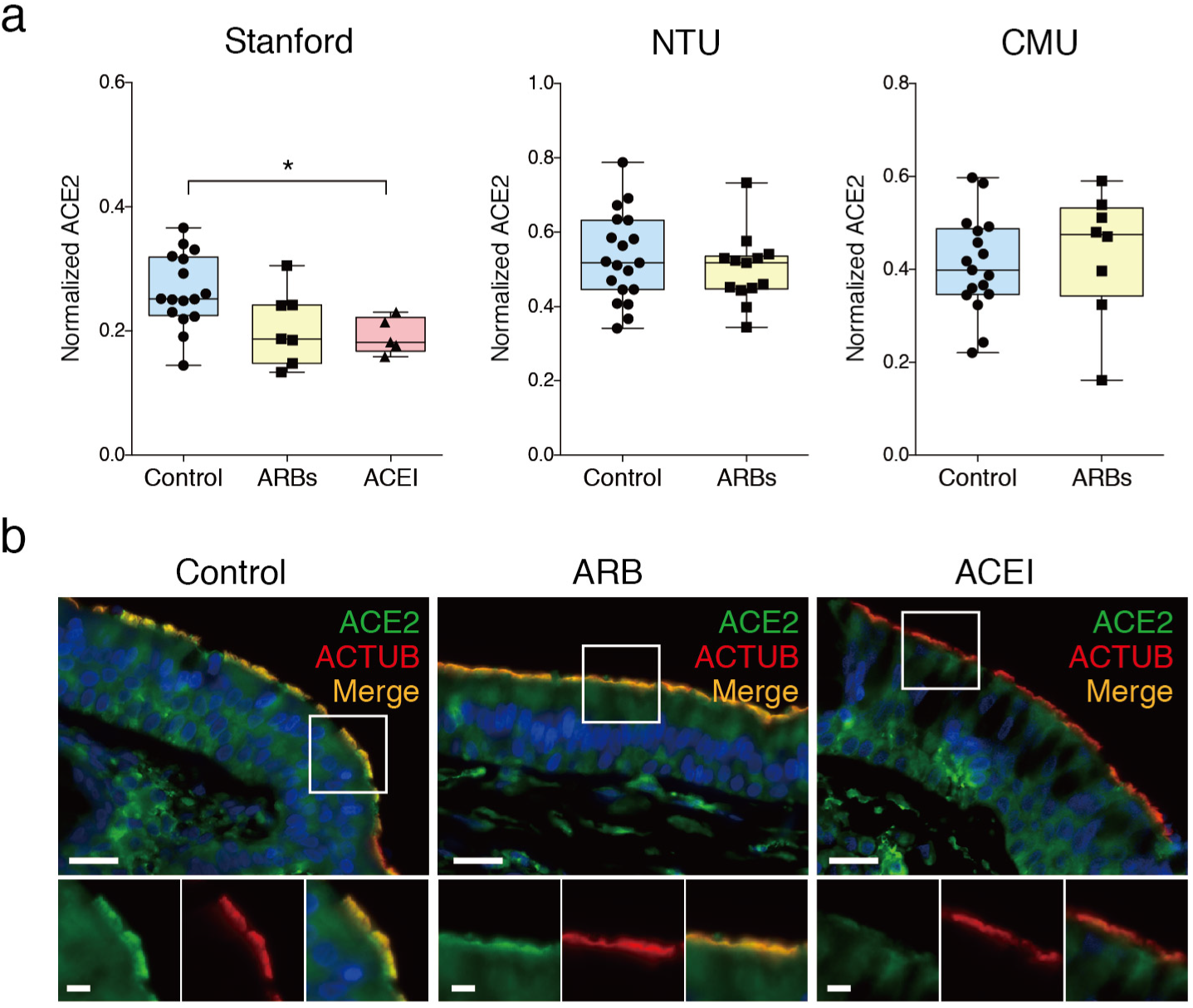
ACE2 expression in the nasal cilia is not increased in patients taking angiotensin-converting enzyme inhibitors (ACEI) or angiotensin II receptor blockers (ARBs), but potentially decreased in ACEI patients. **a**, Quantification of ACE2 in controls and patients taking ARB and ACEI. In the Stanford cohort, ACE2 is slightly but statistically significantly lower in patients taking ACEI (0.19 ± 0.03) compared to controls (0.26 ± 0.06). (Kruskal-Wallis test p=0.012; Dunn’s multiple comparison post-hoc test, *p< 0.05). There were no statistically significant differences in ACE2 expression between patients taking ARB and controls in the Stanford, National Taiwan University (NTU), and China Medical University (CMU) cohorts. All data are noted as mean ± SD. **b**, Representative images displaying the slight but significantly decreased ACE2 in patients taking ACEI, and the non-significant trend towards lower ACE2 expression in the cilia (marked by ACTUB) of patients taking ARB. Scale bars: 20 μm (top); 5 μm (bottom).

**Extended Data Fig. 6.**
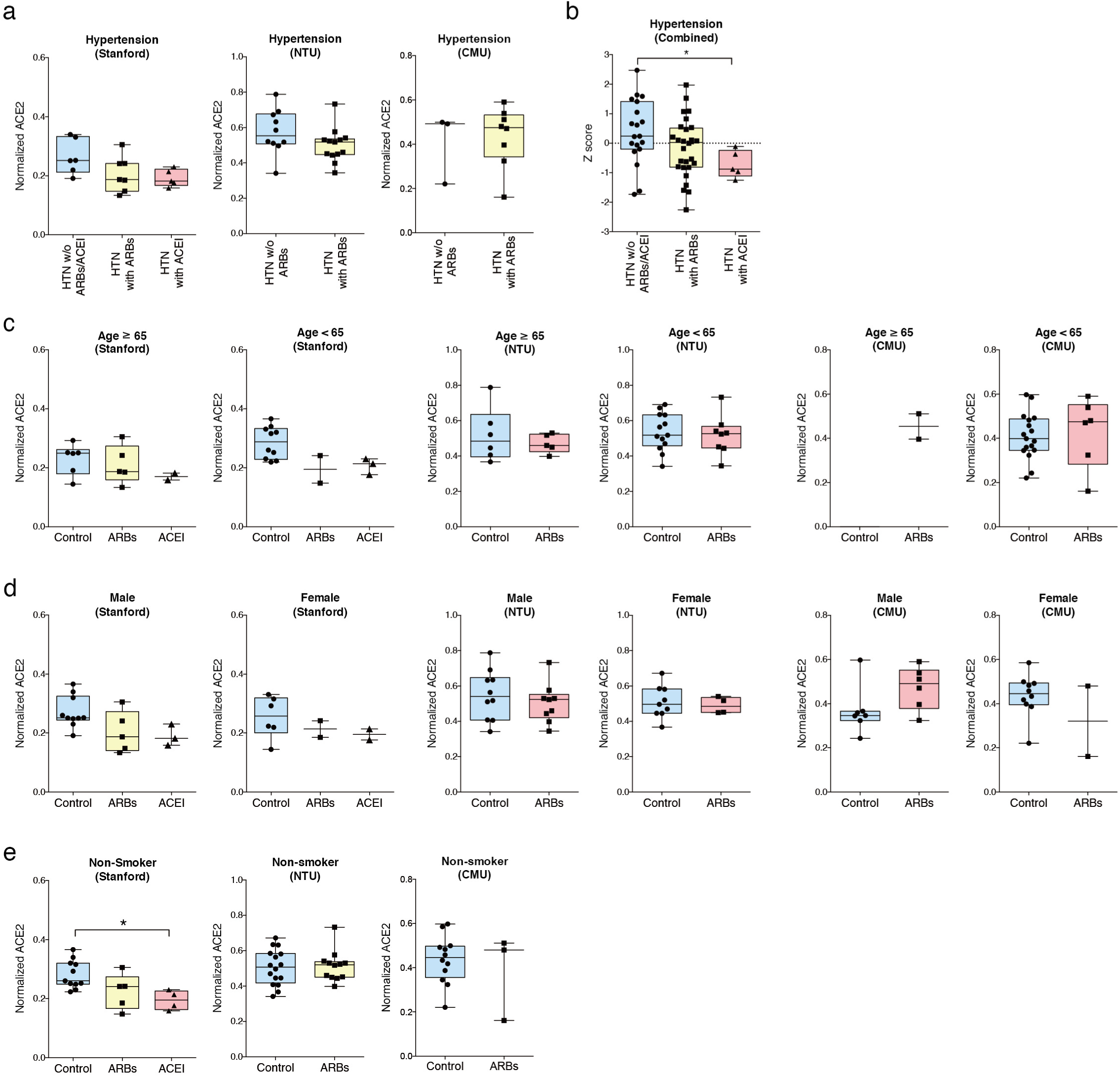
Subgroup analysis of ciliary ACE2 expression among patients taking ARBs or ACEI. **a**, In the Stanford cohort, when including only controls with hypertension (HTN) on other medications (“HTN w/o ARBs/ACEI”), ACE2 expression was statistically different between the groups (Kruskal-Wallis test, p=0.048) but Dunn’s multiple comparison post-hoc test did not reveal any statistical significance between the three groups. **b**, When cohorts from all three institutions were normalized by Z score and integrated, patients taking ACEI (−0.71 ± 0.46) had a lower ACE2 expression compared to controls with hypertension (0.40 ± 1.09). (Kruskal-Wallis test, p=0.035; Dunn’s multiple comparison post-hoc test, *p<0.05). Patients taking ARBs (−0.16 ± 0.99) showed a trend towards lower ACE2 compared to controls with hypertension, but this was not statistically significant. **c**, ACE2 expression among patients of older age (≥65 years old) and patients <65 years old taking ARBs and ACEI was not statistically divergent from control patients of the same age group. (Kruskal-Wallis test, p>0.05). **d**, ACE2 expression among male and female patients on ARBs or ACEI trended lower than same-sex controls except for males taking ARBs in the CMU group who showed a trend towards higher ACE2 expression. No statistically significant differences were observed. (Kruskal-Wallis test, p>0.05). **e**, Among non-smokers, there was a statistically significant trend towards lower ACE2 expression in patients taking ACEI compared to controls in the Stanford group (Kruskal-Wallis test, p=0.005; Dunn’s multiple comparison post-hoc test, *p<0.05). No statistical significance was observed with the non-smokers on ARBs.

The clinical portion of our study has several limitations. First, sample sizes in the treatment groups, especially the ACEI group, were small, increasing the possibility of a Type II error. However, our null hypothesis was that ACEI/ARBs treatment would *increase* ACE2 expression. Instead, we observe a *decrease* in ACE2 among the treatment groups (with the ACEI group reaching statistical significance). The true probability that ACE2 is increased following ACEI or ARBs treatment is therefore likely low. Second, recall bias may be present as information on ACEI/ARBs usage was collected retrospectively, although this was minimized by including patients with confirmed long-term ACEI/ARBs by direct interview and through the electronic medical or pharmacy record. ACEI and ARBs are also long-term medications that are less likely to be used intermittently. Lastly, this was an observational study in which the use of antihypertensive agents for each patient was not randomized, and therefore the ability to make a causal inference is limited due to confounding factors. Although we minimized these elements by using controls matched for age, sex, and smoking status, there are likely other yet unidentified factors for SARS-CoV-2 infection. Clinical trials (NCT04338009 and NCT04312009) are currently underway and will further ascertain the impact of continuation versus discontinuation of ACEI and ARBs on outcomes in patients with COVID-19.

In conclusion, we find that ACE2 protein expression is not only present in ciliated epithelial cells in the human respiratory tract, but that on a subcellular level, it is enriched in the motile cilia organelle of the ciliated epithelium. The latter is anatomically the probable initial or early subcellular site of SARS-CoV-2 viral entry in these cells (and perhaps other cell types). The identification of ACE2 localization to the cilia will guide future functional studies and provide potential mechanisms and targets for strategies that modulate SARS-CoV-2 viral entry and infection routes. The endogenous function of ACE2 in the cilia may be distinct from its role in the RAAS, remains to be determined, and begs the question as to whether SARS-CoV-2, through interaction with ACE2, might interfere with ciliary function during late viremia. Finally, these results support the conclusion of several professional medical societies that have endorsed maintaining standard ACE inhibitors and ARBs therapy during the ongoing SARS-CoV-2 outbreak, although those prior recommendations were provided based on the absence of evidence that ACE2 levels may be increased by ACEI/ARBs. Here, we show the first expression-based evidence that ACE2 protein levels in the respiratory tract are indeed not increased in patients taking long-term ACEI and ARBs, suggesting that these medications can be safely continued as standard antihypertensive therapeutics.

## Methods

### Human nasal tissue specimen collection

Tissues from the nasal cavity and the paranasal sinuses were collected from both healthy control donors and patients with chronic rhinosinusitis from 2018-2020 at the Stanford Sinus Center, National Taiwan University (NTU), and China Medical University (CMU) in Taiwan. Controls represented patients without history, endoscopic, or radiographic evidence of sinus disease, but underwent sinus procedures for surgical access such as for repair of cerebrospinal fluid leaks. Detailed patient characteristics including demographics, medical history, and past medication use were collected in parallel with tissue sample acquisition. Patient data, including medication history, were independently verified through direct interview by a research technician/physician and by a questionnaire additionally administered on the day of surgery to confirm accuracy of existing records from patients’ electronic medical or pharmacy records. Samples were included if the use or non-use of ACEI or ARBs could be confirmed in-person and by electronic medical or pharmacy records. All tissue specimens and patient medical record information, including demographics, medical/social history, and medications were collected under approved Institutional Review Board (IRB) protocols in accordance with the regulations of the Research Compliance Office at the respective institutions. Following surgical excision, nasal specimens were placed in physiologic saline, immediately transported to the lab, and placed in 10% neutral buffered formalin for 24-48 hours before paraffin embedding. Nasal turbinate, uncinate process, and ethmoid sinus tissues were placed into EDTA for bone decalcification prior to embedding into tissue blocks.

### Human lung, bronchial, tracheal tissues

Formalin-fixed, paraffin-embedded (FFPE) tissue blocks from Stanford Pathology archives were selected based on normal histology using a hematoxylin-eosin (H&E) stained tissue section for evaluation. Normal histology was reconfirmed by a board-certified pathologist in the Nolan lab.

### IMCD3 cell culture and transfection

IMCD3 cells were grown in DMEM (11995065, Thermo Fisher Scientific) supplemented with 10% FBS (100-106, Gemini), GlutaMax supplement (35050-079, Thermo Fisher Scientific), 100 U/mL Penicillin-Streptomycin (15140163, Thermo Fisher Scientific) at 37°C in 5% CO2. For transient transfection of IMCD3 cells, IMCD3 cells were grown to ~80% confluence and transfected with 5 μg of *ACE2* plasmid DNA/1×10^6^ cells using Fugene6 (Promega).

### Immunofluorescence immunohistochemistry and imaging

Sections were cut to 4 μm thickness at the Stanford University Histology Service Center and mounted on frosted glass slides. H&E stained sections were obtained from each FFPE block. Deparaffinization, rehydration, and Heat-Induced Epitope Retrieval (HIER) were performed on an a ST4020 small linear stainer (Leica). For deparaffinization, slides were baked at 70°C for 1-1.5 h, followed by rehydration in descending concentrations of ethanol (100% twice, 95% twice, 80%, 70%, ddH2O twice; each step for 30 seconds). Washes were performed using a Leica ST4020 Linear Stainer (Leica Biosystems, Wetzlar, Germany) programmed to 3 dips per wash for 30 s each. Heat-induced epitope retrieval (HIER) was performed in a Lab VisionTM PT module (Thermo Fisher) using Dako Target Retrieval Solution, pH 9 (S236784-2, DAKO Agilent) at 97 °C for 10 min and cooled down to 65°C. After further cooling to room temperature for 30 min, slides were washed for 10 min 3 times in Tris-Buffered Saline (TBS), containing 0.1% Tween® 20 (Cell Marque; TBS-T). Sections were then blocked in 5% normal donkey serum in TBS-T at room temperature for 1 h, followed by incubation with primary antibodies in the blocking solution. After 1 overnight incubation of primary antibodies in 4°C, sections were washed 3 times with TBS-T and stained with the appropriate secondary antibodies in PBS with 3% bovine serum albumin, 0.4% saponin, and 0.02% sodium azide at room temperature for 1 h. Following this, sections were washed 3 times with TBS-T and mounted with ProLong® Gold Antifade mounting medium with DAPI (Invitrogen). The primary antibodies and final titrations used were rabbit anti-ACE2 (1:100; Abcam ab15348), rabbit anti-ACE2 (1:200; Sigma HPA000288), rabbit anti-ACE2 (1:100; Abcam ab239924), rabbit anti-ACE2 (1:100; Novus NBP2-67692), goat anti-ACE2 (1:100; R&D Systems AF933), mouse anti-acetylated α Tubulin (1:300; Santa Cruz sc-23950), mouse anti-MUC-1 (1:100; NSJ Bio V2372SAF), mouse anti-MUC5AC (1:200; Abcam ab212636); mouse anti-CD31 (1:300; Novus NBP2-47785); rabbit IgG isotype control (same mcg as control; Abcam ab172730). Secondary antibodies include donkey anti-rabbit Alexa Fluor Plus 647 1:500 (Invitrogen), donkey anti-mouse Alexa Fluor Plus 555 1:500 (Invitrogen). Fluorescence-immunolabeled images were acquired using a Zeiss AxioImager Z1 microscope. Post-imaging processing was performed using ImageJ. Figures were organized using Adobe Illustrator.

### Chromogenic immunohistochemistry

After deparaffinization and rehydration, slides were blocked for endogenous peroxidase in 3% hydrogen peroxide for 15 minutes at room temperature. HIER was performed with Dako Target Retrieval Solution, pH 9 (S236784-2, DAKO Agilent) at 95 °C for 25 min. 2.5% horse serum was used for blocking for 30 minutes at room temperature followed by incubation overnight at 4°C using one of the following primary antibodies: rabbit anti-ACE2 (1:20,000; Abcam ab15348), rabbit anti-ACE2 (1:1,000; Sigma HPA000288), goat anti-ACE2 (1:500; R&D Systems AF933), rabbit anti-ACE2 (1:500; Abcam ab239924), mouse anti-ACE2 (1:200; R&D Systems MAB933), rabbit anti-ACE2 (1:5,000; Novus NBP2-67692). ImmPRESS HRP anti-rabbit, anti-mouse, or anti-goat IgG polymer detection kit (Vector Laboratories, Burlingame, CA) was used as the secondary antibody link for 30 minutes at room temperature. 5 minute Tris/tween buffer washing steps were performed between each incubation step. The immune complexes were visualized with ImmPACT DAB Peroxidase (HRP) Substrate kit (Vector Laboratories). Cell nuclei were counterstained with hematoxylin. Images were scanned and digitized using the Aperio AT2 and viewed using Aperio Imagescope software.

### In situ hybridization (ISH) staining

Tissue sections (4 μm thick) were cut from FFPE tissue blocks and mounted onto glass slides. Slide-tissue sections were baked at 70°C for 1 hour and subsequently soaked in xylene (Thermofisher Scientific) for 10 min × 3 rounds. Rehydration and HIER of tissue sections were performed in a similar manner as described above except that rehydrating washes were done for 180 seconds each. After HIER and cooling, slides were then washed twice with Milli-Q water (Millipore Sigma) and half the section were subjected to a 10 min protease digestion at 40°C with Protease III (322337, Bio-Techne) diluted to 1:20 in 1X PBS. Slides were then washed for 2 × 2 min Milli-Q water before a 15 min H_2_O_2_ block at 40°C (322335, Bio-Techne). Slides were then washed for 2 × 2 min Milli-Q water before an overnight hybridization at 40°C with probes against the human *ACE2* mRNA (848151, Bio-Techne). Amplification of the ISH probes was performed the next day according to manufacturer’s protocol (323100, Bio-Techne), with the final deposition of Cyanine 3 for *ACE2* mRNA probe targets (NEL744001KT, Akoya Biosciences). Slides were then processed exactly as described for IHC staining for anti-MUC5AC (Abcam ab212636) and ACE2 (Abcam ab15348) staining. Fluorescent images were acquired and processed as detailed above.

### Quantification of fluorescence intensity

Samples within each patient cohort were stained simultaneously with rabbit anti-ACE2 (1:100; Abcam ab15348) and mouse anti-acetylated α-Tubulin (ACTUB 1:300; Santa Cruz sc-23950) using the same master mix and identical incubation times under similar staining conditions described above. Isotype controls were stained with rabbit IgG isotype control (Abcam ab172730) and mouse anti-acetylated α Tubulin (1:300; Santa Cruz sc-23950). Exposure times under fluorescence microscopy were identical for samples within the same cohort. Quantitation was performed in the FIJI package of ImageJ open source software. Binary masks were created by thresholding the anti-acetylated α-Tubulin channel using selected cutoff values that produce inclusive outlines of the ACTUB staining. Cellular membranes were segmented (outlined) using continuity of high signal areas (area larger than 1,000 pixels) on binary masks as the criteria. The signal within the membrane areas were computed for both the ACE2 channel and for ACTUB channel. The estimates of ACE2 signal were further corrected by subtracting the average membrane signal observed in isotype control from the average ACE2 channel per membrane measurements. For the sake of cross-sample normalization, the ratio of the isotype control-subtracted ACE2 signal divided by the ACTUB signal (“normalized ACE2”) for each patient sample was used for further downstream analysis.

### Statistical analysis

Analyses were performed with IBM SPSS 23 (IBM Corporation, Armonk, NY) and GraphPad Prism 6.0 (GraphPad Software, La Jolla, CA) software. The Student’s t-test was used for 2-group comparisons. Multiple comparisons for intergroup differences were assessed by Kruskal-Wallis one-way analysis of variance, followed by Dunn’s multiple comparison post-hoc test. When integrating data from three institutions, data from each institution were converted to Z-score before applying the above statistical comparison. All data are noted as mean ± SD. A p-value of < 0.05 was considered statistically significant.

## Data Availability

All data generated or analyzed during this study are included in this published article or available upon reasonable request.

## Author Contributions

I.T.L. conceived and coordinated the study. I.T.L., T.N., C-T.W., S.J., P.A.G. designed and performed the experiments. C-T.W. performed the microscopy imaging. I.T.L., T.N., P.A.G., C-K.L., L-C.S., C.M.S., D.R.M., P.C., N.A.B., D.Z, S.S.D., A.Y., D.K., K.M.P., R.K., J.B.O., M.A.T., C.H.Y., Y-T.L, C-F. L., D-T.B., G.J.T., Z.M.P., Y-A.T., C-J.T., T-H.Y., P.H.H. consented patients, collected, processed, banked, and/or evaluated the human samples. I.T.L., T.N., C-T.W., Y.G., S.J. analyzed the data. Y.G. developed and performed computational image processing. Y.G. and T.N. conducted statistical analyses. T.N. and C-T.W. prepared the final figures. I.T.L. wrote the manuscript with contributions by T.N., C-T.W., Y.G., S.J., C.M.S., D.R.M., P.C., G.P.N., J.V.N., P.K.J. Funding and supervision were provided by G.P.N., J.V.N., and P.K.J.

## Acknowledgments

We thank members of the Nolan, Nayak, Jackson, Yeh, Tsay, and Stanford Pathology laboratories for helpful discussions and technical assistance. We thank Polly Kavanaugh, Nicole Wang, Carol Valencia, Rebekah Youkhana, Alfred Machicado, Jason Irwin, Camilla Morrison, and Yuka Lee for excellent laboratory and administrative support. We are grateful to faculties of the Stanford Allergy/Immunology Division, especially Drs. Dave Lewis, Yael Gernez, and Sean McGhee for informal guidance and accommodations made for Ivan T. Lee’s protected research time. We thank Mark Kittisopikul for informal discussion on statistical analysis. This work was supported by the Parker Institute for Cancer Immunotherapy (G.P.N.), Food and Drug Administration (HHSF223201610018C and DSTL/AGR00980) (G.P.N.), Fast Grant Funding for COVID-19 Science (G.P.N. and P.K.J.), the National Institutes of Health 1R01AI149672-01 (G.P.N.), P30DK116074 (P.K.J.), the Rachford & Carlotta A. Harris Endowed Chair (G.P.N.), California Institute for Regenerative Medicine (DISC2-09637) (J.V.N.), Defense Advanced Research Project Agency (HR001118S0037-PREPARE-FP-001) (J.V.N.), The Stanford Initiative to Cure Hearing Loss (SICHL) (P.A.G., J.V.N.), The Operndorf Foundation (P.A.G., J.V.N.), The PD Foundation (J.V.N.), Stanford Translational Research and Applied Medicine (TRAM) Pilot Grant (I.T.L.), Thrasher Research Fund Early Career Award (I.T.L.), Stanford Maternal and Child Health Research Institute (MCHRI) Clinical (MD) Trainee Support Award (I.T.L., Ernest and Amelia Gallo Endowed Postdoctoral Fellow), Leukemia & Lymphoma Society Career Development Program (S.J.).

